# Gut Microbiome Multi-Omics and Cognitive Function in the Hispanic Community Health Study/Study of Latinos- Investigation of Neurocognitive Aging

**DOI:** 10.1101/2024.05.17.24307533

**Authors:** Natalia Palacios, Scott Gordon, Tao Wang, Robert Burk, Qibin Qi, Curtis Huttenhower, Hector M. Gonzalez, Robert Knight, Charles De Carli, Marta Daviglus, Melissa Lamar, Gregory Telavera, Wassim Tarraf, Tomasz Kosciolek, Jianwen Cai, Robert C. Kaplan

**Affiliations:** Department of Public Health, University of Massachusetts Lowell, Lowell, MA, 01850; Bedford VA Healthcare System, Geriatric Research and Education Clinical Center, Bedford, Massachusetts, USA, 01730; Harvard Chan Microbiome in Public Health Center (HCMPH), Boston, MA, 02115; Albert Einstein College of Medicine, Bronx, NY, 10461; Broad Institute of MIT and Harvard, Cambridge, MA, 02142; Department of Immunology and Infectious Diseases, Harvard T.H. Chan School of Public Health, Boston, MA, 02115; Department of Biostatistics, Harvard T.H. Chan School of Public Health, Boston, MA, 02115; University of California, San Diego Department of Neurosciences, San Diego, CA, USA 97037; Department of Bioengineering, University of California, San Diego, La Jolla, CA, USA, 97037; Department of Pediatrics, University of California, San Diego, La Jolla, CA, USA, 97037; Center for Microbiome Innovation, University of California, San Diego, La Jolla, CA, USA, 97037; University of California, Davis, CA, USA, 95616; University of Illinois College of Medicine Institute for Minority Health Research, Chicago, IL, 60612; Rush University Alzheimer’s Disease Center, Rush University, Chicago, IL, 60612; South Bay Latino Research Center, San Diego State University, Chula Vista, CA, 92182; Wayne State University, Detroit, MI, 48202; Sano Centre for Computational Precision Medicine, Krakow, Poland; Department of Biostatistics, University of North Carolina at Chapel Hill, Chapel Hill, NC, 27599

**Author notes:** Corresponding author: Natalia Palacios, Department of Public Health, University of Massachusetts, Lowell.

**Keywords:** Hispanic, Latino, Cognition, Microbiome, Metabolomics

## Abstract

**INTRODUCTION:** *We* conducted a study within the Hispanic Community Health Study/Study of Latinos- Investigation of Neurocognitive Aging (HCHS/SOL-INCA) cohort to examine the association between gut microbiome and cognitive function.

**METHODS:** We analyzed the fecal metagenomes of 2,471 HCHS/SOL-INCA participants to, cross-sectionally, identify microbial taxonomic and functional features associated with global cognitive function. Omnibus (PERMANOVA) and feature-wise analyses (MaAsLin2) were conducted to identify microbiome-cognition associations, and specific microbial species and pathways (Kyoto Encyclopedia of Genes and Genomes (KEGG modules) associated with cognition.

**RESULTS:** *Eubacterium* species(*E. siraeum* and *E. eligens*), were associated with better cognition. Several KEGG modules, most strongly Ornithine, Serine biosynthesis and Urea Cycle, were associated with worse cognition.

**DISCUSSION:** In a large Hispanic/Latino cohort, we identified several microbial taxa and KEGG pathways associated with cognition.

## BACKGROUND

The human gut microbiome comprises trillions of bacteria, more microorganisms than the number of cells in a human body and 150 times as many genes as the human genome[1]. The microbiome plays key roles in absorption and metabolism of nutrients, breakdown of lipids and polysaccharides, detoxification of xenobiotics, waste particles and pathogenic organisms[2, 3], control of gut motility and homeostasis of the intestinal barrier[4, 5]. It is key to the development and function of the human immune system, with 70% of the body’s lymphocytes located in the gut[6, 7]. The intestinal environment is intimately connected to the central nervous system (CNS) through bi-directional communication pathways regulated, in part, by the gut microbiome via production of bioactive metabolites[8, 9]: the brain-gut-microbiome axis. This communication system is just beginning to be appreciated and includes neural, immune,[10] endocrine, and metabolic pathways[9].

US Hispanics/Latinos are at an increased risk of AD/ADRD and accelerated cognitive decline compared to non-Hispanic/Latino whites[11–14]. Hispanics/Latinos are also one of the fastest growing segments of the US elderly population[15, 16], and are impacted by cognitive decline at younger ages[17–19] compared to non-Hispanic Whites. This elevated risk has been suggested to be due to attributed to higher prevalence of key AD/ADRD risk factors in Hispanics/Latinos, such as poor diet, higher prevalence of diabetes, metabolic syndrome, obesity and cardiovascular disease, and higher blood pressure, which have been documented in HCHS/SOL-INCA [11, 20–27]. A distinct microbial profile in Hispanics/Latinos may also contribute to increased risk[28].

Gut inflammation has been associated with AD/ADRD risk[29], suggesting that the gut microbiome may contribute to ‘inflammaging’ in AD/ADRD. Prior studies focusing on microbiome and AD/ADRD have reported reduction in diversity[30], and lower abundance of beneficial anti-inflammatory taxa, such as *Eubacterium spp.*[31] and *Feacalibacterium spp.*[32, 33] in AD/ADRD. Several species of gut bacteria, such as *Bifidobacterium*, are capable of production of neuroactive metabolites, such as serotonin and GABA.[34–36] Several bacteria including the *Klebsiella, Escherichia, Streptococcus, and Salmonella*, *Pseudomonas*, species secrete functional amyloid proteins with demonstrated capacity to cross-seed and trigger a cascade of amyloid protein misfolding[37, 38] that can propagate from the gastrointestinal tract to the brain. These amyloid-producing taxa have been reported elevated in AD/ADRD in prior studies[39–41]. Furthermore, secretion of lipopolysaccharide (LPS), a cell wall component of gram negative bacteria[42], can lead to microglial priming, increased production of proinflammatory cytokines, neuroinflammation and neurodegeneration[43]. Pathogens, such as *K. pneumoniae and E. lenta* [44, 45], and pro-inflammatory taxa, such as the sulfate-producing *Desulfovibrio*have been noted in patients with AD/ADRD.

Functional work on the microbiome and AD has implicated both inflammatory and metabolic processes in AD. For example, finding of dysregulation of the P-glycoprotein microbial pathway[46] in AD, suggests a contribution of intestinal inflammation and gastrointestinal infections. Other studies reported dysregulation in pathways related to glucose metabolism and mitochondrial disfunction[47].

Animal studies have supported this epidemiological evidence in humans, reporting altered microbial composition in transgenic AD mouse models[48], and absence of amyloid plaque build-up in germ free mice[49]. Transgenic mice treated with an antibiotic cocktail had fewer insoluble amyloid *β* plaques and less microglia and astrocyte accumulation around existing amyloid plaques[50]. In a recent report based on a fecal microbiome transfer (FMT) from AD patients to microbiota-depleted healthy young adult rats, FMT resulted in AD symptoms in the AD-colonized rats, as well as changes in the rat cecal and hippocampal metabolomes[51]. The abundance of the pathobiont, pro-inflammatory *Desulfovibrio* was elevated in AD patients compared to healthy controls, inversely correlated to MMSE scores and was the taxa most elevated in the AD-colonized rats after FMT[51]. *Desulfovibrio* has been reported enriched in AD patients by several other studies[46, 52] and has been associated with reduced cecal SCFA levels[53].

To knowledge, no work on the gut microbiome and AD risk has been done in Hispanics/Latinos. We conducted the first study of the gut microbiome in relation to cognitive function in a Hispanics/Latinos cohort.

## METHODS

The study was approved by the institutional review boards at Albert Einstein College of Medicine and the University of Massachusetts at Lowell.

### Study Population

The study was conducted within Hispanic Community Health Study/Study of Latinos (HCHS/SOL) a large population-based, multisite, prospective cohort study cohort of US Hispanic adults supported by the National Heart Lung and Blood Institute (NHLBI) and other National Institutes of Health (NIH) institutes. Participants (N=16,415) were enrolled during 2008-2011 at four Field Centers located in US cities with large Hispanic/Latino populations: Bronx, NY; Chicago, IL; Miami, FL; and San Diego, CA. Participants are of Cuban, Dominican, Puerto Rican, Mexican, Central and South American backgrounds. Middle-aged and older Hispanics/Latinos (ages 45-74 years) were oversampled (n=9,652). At study baseline, participants participated in in-clinic visits with bilingual technicians that included anthropometric measures, blood pressure readings, pulmonary testing, diet assessment as well as a blood draw (with blood frozen and stored) [54, 55].

### Neurocognitive assessment

At the second HCHS/SOL visit (V2), neurocognitive testing was conducted among 6,377 participants, at the 2^nd^ HCHS/SOL study visit completed during 2011-2014. All cognitive assessments were performed in-clinic by trained bilingual/bicultural technicians with oversight of a Neurocognitive Reading Center. All three waves included Six-Item Screener[56] (SIS; mental status), (2) Brief-Spanish English Verbal Learning Test (BSEVLT; verbal episodic learning and memory),[57] (3) Verbal Fluency[58] and (4) Digit Symbol Subtest (DSS; processing speed).[59] At v2 and v3, additional testing included the Trail Making Test (TMT; A&B; a test of cognitive function), the NIH Toolbox Picture Vocabulary Test (PVT; test of general cognitive ability/crystalized knowledge[60] and the 12-item Everyday Cognition (eCog-12) scale of (memory, language, visuospatial, planning, organization, and divided attention)[61]. In this study, our cognitive outcome variable was global cognition scores (GSC), derived from confirmatory factor analyses based on the cognitive measures above[62].

### Stool Sample Collection and Storage

During HCHS/SOL V2, participants were provided with a stool collection kit for self-sampling using a disposable paper inverted hat (Protocult collection device, ABC Medical Enterprises, Inc., Rochester, MN). CHS/SOL collected and sequenced over 3,035 stool samples during the V2 phase, placed in RNAlater[28]. Of these 2,470, were part of HCHS/SOL-INCA and had GCS, stool metagenomic and were included in this study.

### Fecal sample processing and sequencing

Shotgun sequencing was performed in the laboratory of Dr. Knight, using previously described procedures[63]. Briefly, DNA was extracted from fecal samples on FTA cards, following the Earth Microbiome Project protocol[64, 65]. Input DNA was quantified, and thereafter DNA fragmentation is performed, followed by end-repair and A-tailing. Adapters and barcode indices are added following the iTru adapter protocol[66]. Each plate of 384 libraries is generated without repeating barcodes, eliminating the problem of sequence misassignment due to barcode swapping[67]. The libraries were purified, quantified and normalized for sequencing on Illumina NovaSeq. Sequencing depth ranged from 500K-8,945K reads/sample, with average 955K reads/sample.

### Bioinformatic profiling of fecal samples

FASTQ sequence reads were demultiplexed, filtered to remove reads mapping to the human genome, and trimmed to remove low quality bases. The reads are then aligned against the NCBI RefSeq representative prokaryotic genome collection (release 82)[68] using Bowtie2,[69] and per strain coverage calculated using default SHOGUN[63] settings. The α-diversity indices (Shannon) and β-diversity (Bray-Curtis dissimilarity) were calculated using R vegan packages[70, 71]. Functional profiles were obtained using SHOGUN via sequence alignment to a nucleotide gene database derived from NCBI RefSeq (release 82) and annotated with Kyoto Encyclopedia of Genes and Genomes (KEGG) orthology[72, 73].

### Metabolomic Profiling

Metabolomics data, capturing >600 metabolites within eight defined classes, was obtained using fasting plasma specimens collected at HCHS/SOL V2. Metabolomics profiling was conducted via Metabolon HD4Discovery platform (Metabolon Inc., Durham, NC) (Morrisville, NC) using liquid chromatography-MS/MS methods with positive ion and negative ion modes (Waters ACQUITY ultra-performance liquid chromatography, as previously described[74]. Of the 2,471 participants with metagenomic and cognitive data at V2, 480 had concurrent serum metabolome data. Data on these 2 participants were used in multi-omic analyses below.

### Assessment of Covariates

Comprehensive covariate data were collected during in-person examinations, as well as annual telephone interviews and ongoing ascertainment of major incident health events. Our model was adjusted for factors previously associated with AD, such as age, biological sex, education, AHEI2010 dietary index, metformin use, Hispanic background (Central American, South American, Dominican, Mexican, Cuban, Puerto Rican), age moved to US, fiber intake and diabetes status. Age, education, metformin use[75], Hispanic background and years living in the US[28] were collected as part of in-person interview at V2. Definition of diabetes and prediabetes are based on contemporaneous ADA criteria by a combination of diagnosis and measured levels of fasting and 2hr glucose and hemoglobin A1c tests.[76] Long-term diet is one of the primary factors driving human microbiome composition and function[77], including among immigrants[78, 79]. In HCHS/SOL, all participants completed, at baseline, two 24h dietary recalls at ∼6 week intervals, to capture usual intake of foods and nutrients[80], from which fiber use and Alternative Healthy Eating/AHEI score were derived. We adjusted our analyses for use of Metformin use, as it is used by ∼12% of our participants, is the most common diabetes medication, has been shown to lead to alterations in gut microbiome composition and function.[81] We adjusted for Hispanic background and, in sensitivity analyses, for years living in the US as these factors were strongly related to the microbiome of HCHS/SOL participants in our prior studies.

### Statistical Analysis

All analyses were conducted on 2,471 HCHS/SOL-INCA participants who had concurrent GCS and stool metagenomic data (**Figure 1A**). Analyses involving the metabolome, were conducted in the 480 participants who also had serum metabolomic data. Our initial dataset contained 3,314 microbial species after quality control and removal of host ‘contaminant’ sequences, and of these, 198 were retained after filtering (as detailed above) and used in our statistical analyses. Likewise, of 445 KEGG Modules, 216 were retained after filtering (**Figure 1B).**

**Figure 1.**
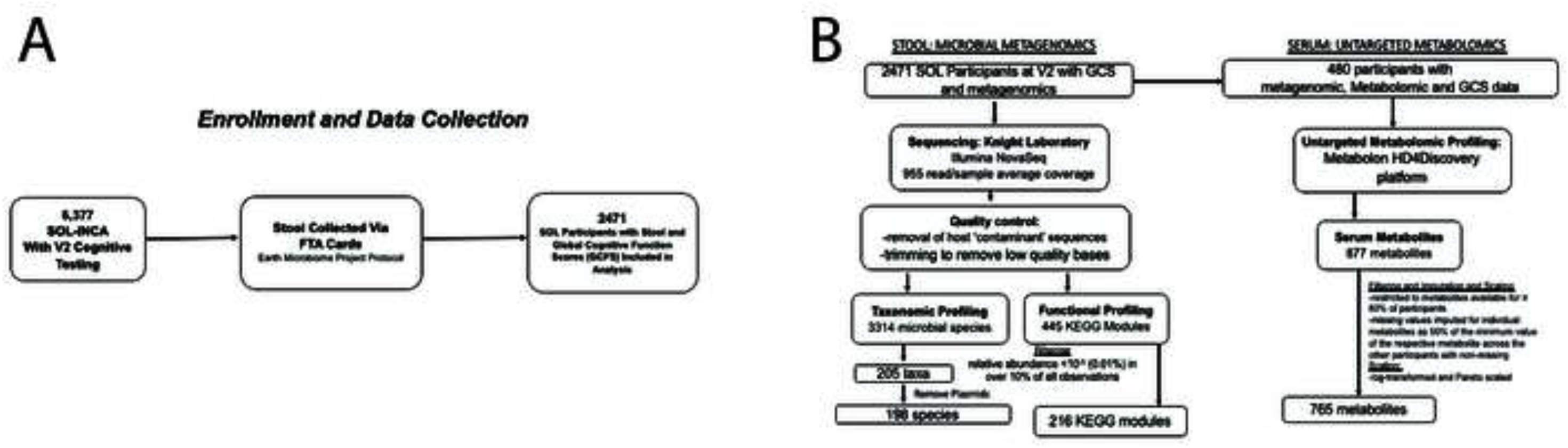
Microbiome and cognition in HCHC/SOL-INCA study design. **Figure 1A**: Sequencing and profiling of HCHC/SOL-INCA Visit 2 samples with microbiome and metabolomics data. **Figure 1B**: STORMS flowchart[125] outlining enrollment, data collection, and processing for our study of human gut metagenomes and metabolomes in relation to global cognitive function.

### Global Cognitive Function Variable and Categories

GCS was treated as a continuous outcome variable in MaAsLin2 and univariate PERMANOVA analyses. For analyses requiring categorical outcomes, including pCoA visualization, GCS was re-categorized into an ordinal variable with the following values: better (>=1SD above mean GCS; N = 413), medium (between 1SD above and 1SD below mean GCS; N = 1621) and worse (>=1SD below mean GCS; N = 429).

### Overall community patterns of microbial variation

The Bray-Curtis dissimilarity metric was used for all beta-diversity analyses, both of taxonomic composition and functional potential. We performed ordination via the Principal Coordinates Analyses (PCoA), to visualize Beta-diversity relationships across the three GCS categories (better, medium, worse cognition). We performed omnibus testing with permutational multivariate analysis of variance (PERMANOVA) of Bray-Curtis dissimilarities to quantify the percent variance explained by GCS (continuous) and study covariates.

### Feature-wise analyses

To identify microbial features and functions associated with GCS, we used Multivariate Association with Linear Models version 2 (MaAsLin2), Version 2.1.16.0, (https://huttenhower.sph.harvard.edu/maaslin2)[82]) a modified general linear model for feature-wise multivariate testing in microbial community profiles. MaAsLin2 was run in R Version 4.3.1. Model specification and covariates were as follows:

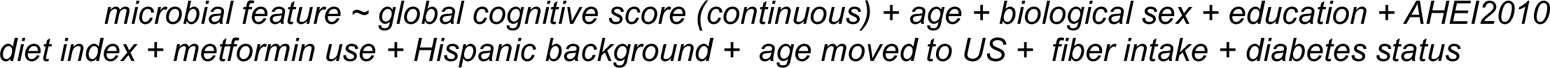

We conducted sensitivity analyses MaAsLin2 including 1) univariate models, 2) additional adjustment for pack years smoking, chronic stress, depression (CESD10), hypertension, waist-hip ratio, alcohol intake, whether participant was US born, and the Multi-Ethnic Study of Atherosclerosis MESA acculturation score[83], a score representing extent of acculturation to the US.

For features (taxa and KEGG modules) most strongly associated with cognition (FDR <= 0.2), we examined the Spearman correlation of these cognition-associated taxa with cognition-associated KEGG modules. To understand the relationship between the identified taxa and KEGG modules to key serum microbial metabolites in serum, we examined the Spearman correlation of these cognition-associated taxa and KEGG modules with serum metabolites from the HCHS/SOL V2 collection. We performed these correlation analyses for serum metabolites within the Short Chain Fatty Acid (SCFA), Branched Chain Amino Acid (BCAA) and tryptophan metabolism networks.

## RESULTS

**Table 1** outlines the characteristics of the study participants with cognitive measures and metagenomic profiles at HCHS/SOL V2 who were included in this study. The average age of study participants was 60.83 years, 10% were US born. The highest proportion of participants (38.46%) were of Mexican heritage, followed by Puerto Rican (18.30%), Cuban (13.85%), Dominican (10.28%), Central American (9.88%), South American (7.04%) and mixed (2.02%) heritage. Most participants were either had pre-diabetes (47%) or diabetes (34%), 12% reported metformin use, and 54.04 had hypertension.

**Table 1.** Characteristics of 2471 HCHS/SOL-INCA participants with cognitive and microbiome measures at HCHS/SOL-INCA V2.

|  | All<br>(N=2471) | Women<br>(N=1625) | Men<br>(N=846) |
| --- | --- | --- | --- |
| Age (mean, SD) | 60.83 (7.11) | 60.77 (7.06) | 60.94 (7.19) |
| Hispanic/Latino Heritage |  |  |  |
| Dominican | 10.28 (254) | 11.51 (187) | 7.92 (67) |
| Central American | 9.88 (244) | 9.98 (162) | 9.69 (82) |
| Cuban | 13.85 (342) | 12.25 (199) | 16.90 (143) |
| Mexican | 38.46 (950) | 40.02 (650) | 35.46 (300) |
| Puerto Rican | 18.30 (452) | 17.36 (282) | 20.09 (170) |
| South American | 7.04 (174) | 68.97 (112) | 7.33 (62) |
| 2+ Heritage | 2.02 (50) | 1.72 (28) | 2.60 (22) |
| Diabetes Status |  |  |  |
| Without Diabetes | 19% | 20% | 18% |
| With Pre-Diabetes | 47% | 47% | 45% |
| With Diabetes | 34% | 33% | 37% |
| MCI at V2 | 9% | 10% | 8% |
| Education: | 33% | 39% | 34% |
| <8 grade |  |  |  |
| 8 grade - HS | 20% | 19% | 20% |
| >HS | 38% | 36% | 41% |
| Born in the US | 10% | 9% | 12% |
| Depression (CESD score) | 7% | 7% | 6% |
| Hypertension | 54.05% (1335) | 53.45% (868) | 55.20% (467) |
| Waist/Hip Ratio | 0.94 (0.085) | 0.92 (0.07) | 0.99 (0.069) |
| Diet/AHEI Score | 51.1 | 50.6 | 52.3 |
| Metformin Use | 12% | 13% | 12% |
| Fiber Intake (g/day) | 19 | 17 | 21 |
| CESD Depression | 6.70 (6.09) | 7.32 (6.30) | 5.53 (5.49) |

### Overall patterns of microbial community variation in relation to GCS

Applying univariate PERMANOVA of Bray-Curtis dissimilarities, we observed variables such as the MESA Acculturation Scale, Sex, Puerto Rican descent and age moved to the US explained the greatest amount of variation in gut microbiome composition (**Figure 2A, Taxa**), while metformin use, age and diabetes explained the largest amount of variation in functional potential (**Figure 2A, KEGG Modules**), consistent with our prior publications[28]. Diet (AHEI dietary score), fiber intake, diabetes, and several other lifestyle variables, also explained significant amount of variation in microbiome composition (**Figure 2A, Taxa**) and function (**Figure 2A, KEGG Modules**). In multivariate PERMANOVA analyses, global cognitive function did not explain a significant amount of variation in microbial taxa (R^2^ = 0.07%; p = 0.22;), or functions (R^2^ = 0.07%; p = 0.24) (**Figure 2B**).

**Figure 2.**
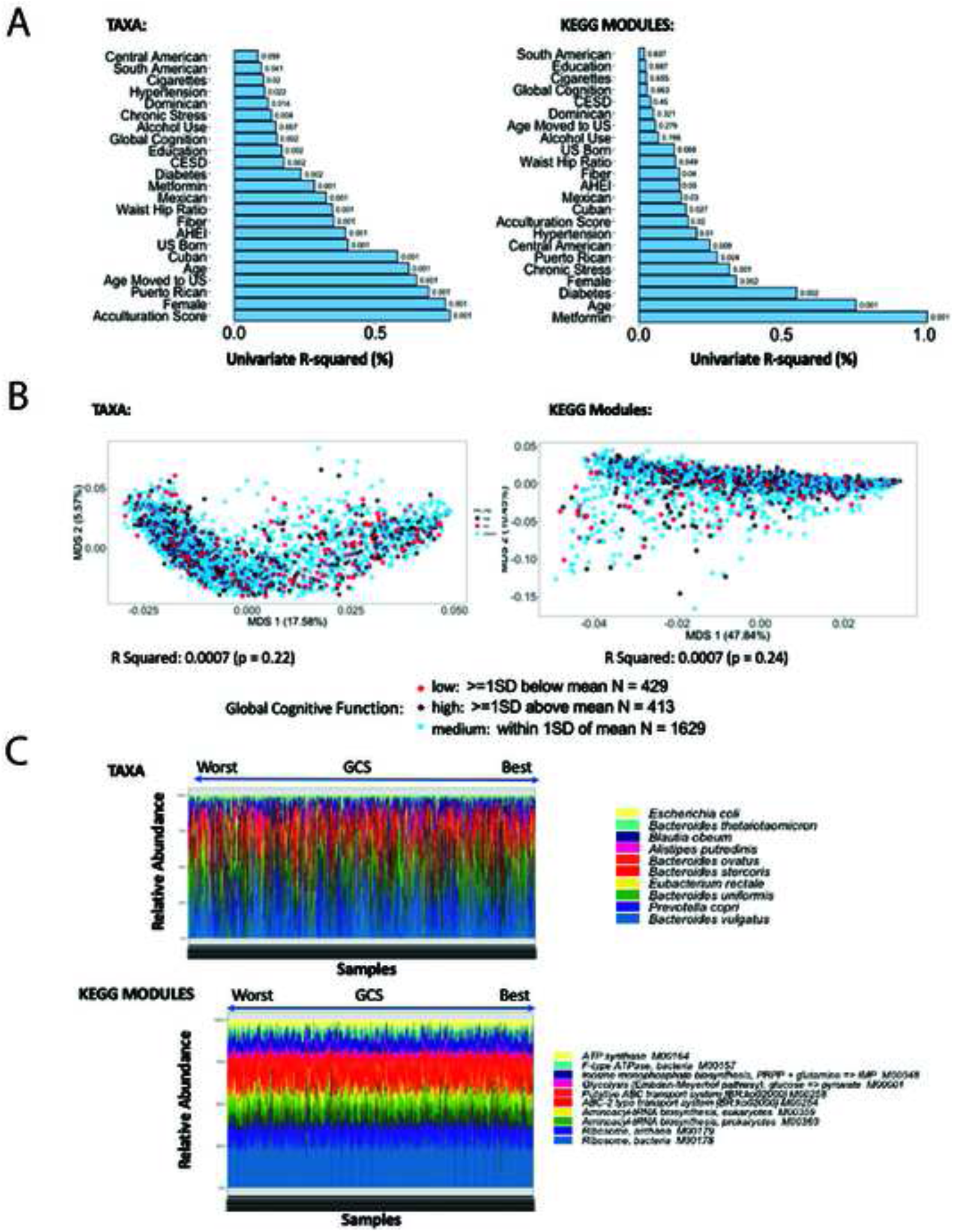
Overall microbiome structure in relation to GCS and other study covariates. **Figure 2A**: Univariate R2 (Bray-Curtis dissimilarities, PERMANOVA) explained by key covariates. **Figure 2B**: Distributions of taxonomic Bray-Curtis dissimilarities by phenotype group show that overall microbiome structure does not vary significantly with global cognitive function. Multivariate PERMANOVA adjusted for adjusted for age, gender, education, diet (AHEI), metformin use, Hispanic background (list). **Figure 2C**: Most abundant Taxa and KEGG Modules in HCHC/SOL-INCA according to GCS.

### Microbial species associated with GCS

The most abundant species in stool included *Bacteroides vulgatus, Prevotella copri, Bacteroides uniformis and Eubacterium rectale* (**Figure 2C, Taxa**). The most abundant KEGG modules included Ribosome bacteria M00178, Ribosome archaea M00179, Aminoacyl-tRNA biosynthesis prokaryotes M00360, Aminoacyl-tRNA biosynthesis eukaryotes M00359 and ABC-2 type transport system (BR:ko02000) M00254 (**Figure 2C, KEGG Modules**).

In feature-wise, MaAsLin2 [84] models, after adjustment for age, biological sex, education, AHEI2010 diet index, metformin use, Hispanic background, age moved to US, fiber intake and diabetes status, we observed an inverse association between several bacterial species, including *Prevotella sp P4.76*, *Bifidobacterium longum Prevotella bryantii, Desulfovibrio piger Ruminococcus faecis*, and GCS. Abundance of several bacterial species, most strongly *Clostridium phoceensis*, *Eubacterium eligens*, and also *Eubacterium siraeum, Holdemania massiliensis*, *Intestimonas massiliensis*, *Bacteroides barnesiae*, and others was positively associated with global cognitive function (**Figure 3A)**. In sensitivity analyses, after additional adjustment for pack years smoking, chronic stress, depression (CESD10), hypertension, waist-hip ratio, alcohol intake, whether participant was US born, and the MESA acculturation score, abundance of *Prevotella sp P4.76* and *Ruminococcus faecis* remained inversely and abundance of *Intestinimonas massiliensis* and *Eubacterium eligens* remained positively associated with GCS (**Supplemental Figure 1**).

**Figure 3.**
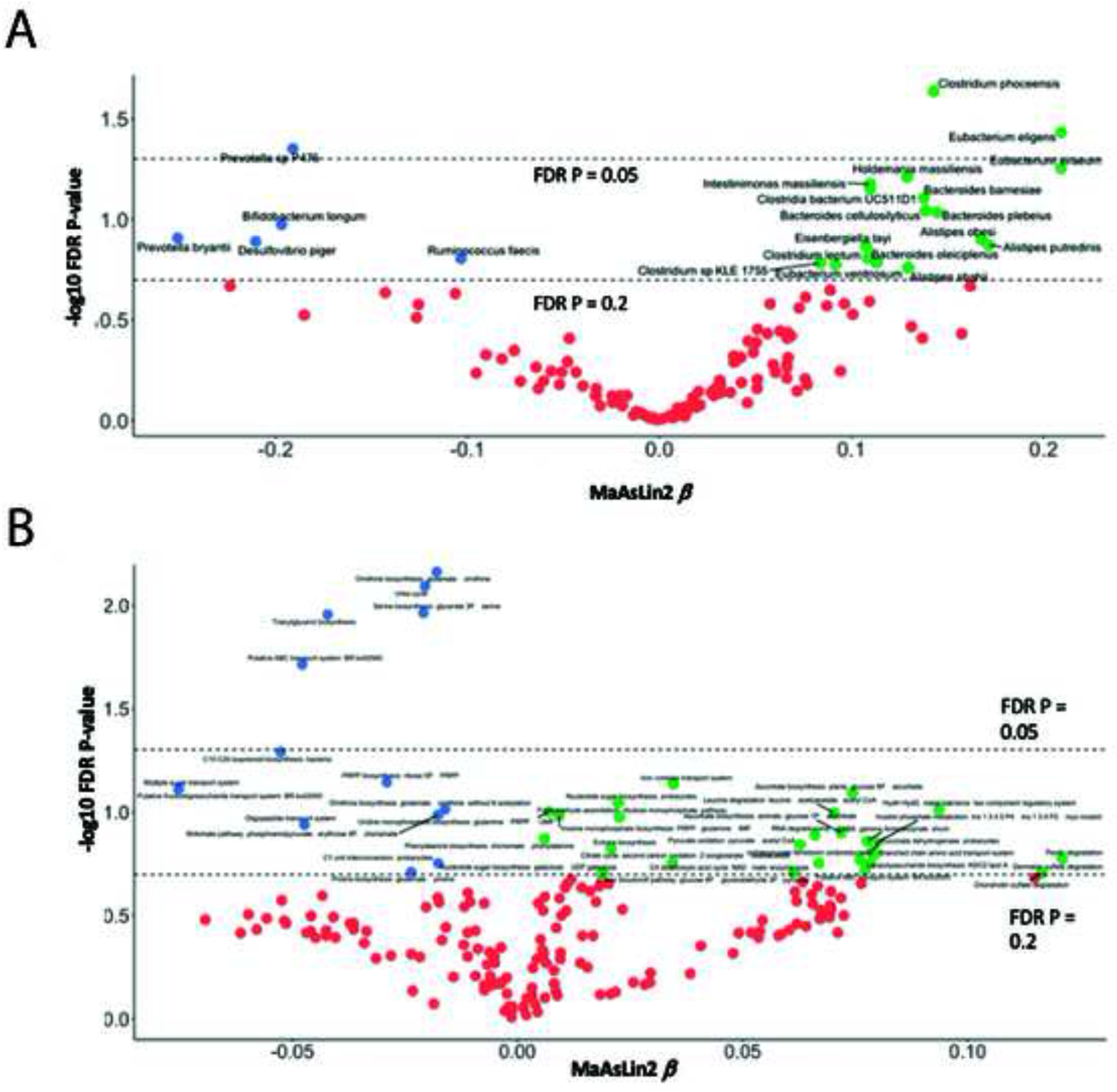
Taxa and KEGG modules significantly associated with GCS. Volcano plots show beta coefficient (x-axis) and FDR-corrected log10(q value) (y-axis). The most significantly enriched and depleted taxa (**Figure 3A**) and KEGG modules (**Figure 3B**) associated with phenotype group identified via feature-wise analyses (MaAsLin 2). All analyses consider global cognitive function as a continuous predictor and are adjusted for age, gender, education, diet (AHEI), metformin use, Hispanic background. All p-values are presented with FDR correction for multiple comparisons.

### Microbial functions associated with GCS

MaAsLin2 identified 14 KEGG Modules significantly associated with GCS (**Figure 3B**). Of these, Ornithine biosynthesis modules (M00536), Urea cycle (M00029), Serine biosynthesis glycerate 3P serine (M00020), Triacylglycerol biosynthesis (M00089), and Putative ABC transport system (M000258), were most strongly associated with worse global cognitive function, with an FDR p<0.05. Abundance of the Iron complex transport system (M003645), Ascorbate biosynthesis plant glucose 6P ascorbate (M00114), nucleotide sugar biosynthesis prokaryotes (M003620, and Leucine degradation leucine acetoacetate acetyl CoA (M00036), among others, modules was positively associated with GCS (**Figure 3B**). In sensitivity analyses, after additional adjustment for pack years smoking, chronic stress, depression (CESD10), hypertension, waist-hip ratio, alcohol intake, whether participant was US born, and the Multi-Ethnic Study of Atherosclerosis MESA acculturation score the KEGG modules that remained inversely associated with GCS at FDR P<0.2) included ornithine biosynthesis glutamate (M00028), serine biosynthesis glycerate 3P serine (M0020), Urea Cycle (M00029) and Putative ABC transport system BR Ko02000 (M00258), no modules were positiviely associated with GCS in these sensitivity analyses. (**Supplemental Figure 2**).

As expected, taxa associated with better cognition were, for the most part, correlated with each other (**Supplemental Figure 2A**). This was also true for taxa associated with worse cognition, with the exception of *B. longum*, the association of which with worse cognition is surprising as it is generally recognized as a beneficial taxon. Thus it’s consistent with prior work that *B.longum* might correlate with beneficial taxa. Similarly, KEGG modules associated with better cognition were positively correlated with each other and negatively correlated with modules associated with rose cognition (**Supplemental Figure 2B).** We observed a modest correlation between, the top taxa and KEGG modules positively associated with cognition, and analogously a correlation between taxa and pathways negatively associated with cognition (**Figure 4A**). *Alistripes shahii* was most strongly correlated to various cognition-associated KEGG modules in this study (Figure 6), with strong correlations with the chondroitin degradation (r = 0.88, FDR P<0.05), sulfate deradation (r = 0.88, FDR P<0.05) and the pectin degradation (r = 0.87, FDR P<0.05) KEGG modules among others. Other strong correlations included the positive correlation between abundance of *Bacteroides plebeius* and HydH HydG metal tolerance two component regulatory system (r=0.45, FDR P <0.05) and the Formaldehyde assimilation ribulose monophosphate module (r = 0.35, FDR P,0.05).

**Figure 4.**
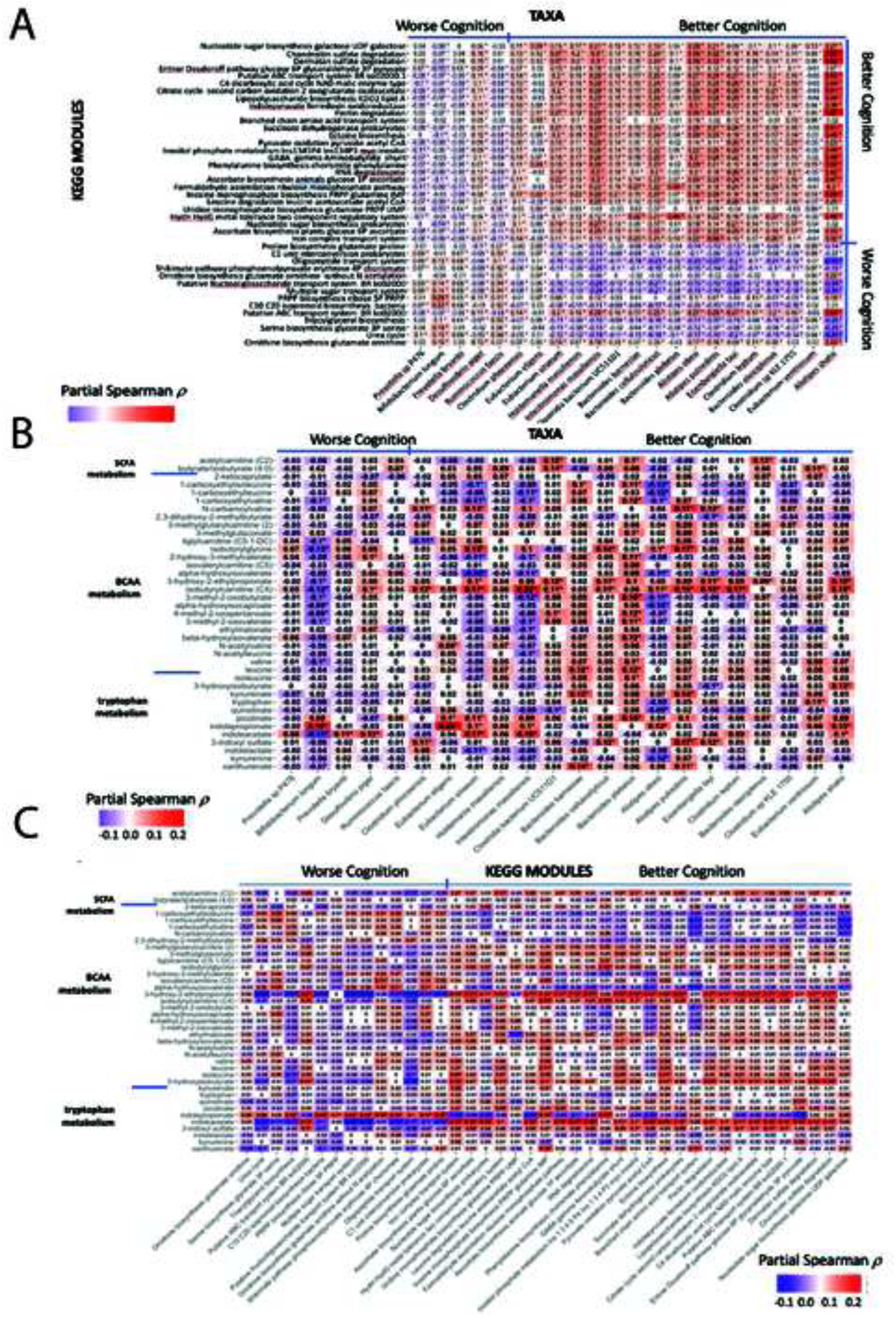
Multi-omic correlations, taxa, KEGG modules and metabolites. Correlation coefficients are partial Spearman correlations adjusted for age, gender, education, diet (AHEI), metformin use, Hispanic background. **Figure 4A**: We observed modest, but consistent correlation between top cognition-associated taxa and KEGG modules (FDR <0.2). Cognition-positive taxa were generally correlated with cognition-positive pathways and, likewise, cognition-negative taxa were overall associated with cognition-negative pathways. **Figure 4B**: Correlation between top cognition-associated taxa (FDR <0.2) with and metabolites in the SCFA, BCAA and tryptophan pathways in HCHS/SOL. **Figure 4C**: Correlation between top cognition-associated KEGG modules (FDR <0.2) with and metabolites in the SCFA, BCAA and tryptophan pathways in HCHS/SOL-INCA.

Several cognition-associated taxa identified in this study were correlated with concurrently collected (HCHS/SOL V2) serum metabolites within SCFA, BCAA and tryptophan metabolic networks. Consistent with results previously published by our group[85], *B. longum,* a beneficial taxa surprisingly associated with worse cognition in our study, was negatively correlated to most metabolites in the BCAA pathway, and strongly positively correlated with indolepropionate (r = 0.19, FDR P<0.05). *B. longum* was also inversely correlated with most BCAA metabolites. We also observed, a positive correlation between indolepriopionate and many of taxa positively associated with cognition, including *E. eligens* (r = 0.28, FDR P,0.05) and *E. siraeum* (r = 0.1, FDR P<0.05), and *I. massiliensis* (r=0.1, FDR p<0.05), *A.obesi* (r = 0.12, FDR P<0.05) and *A. shahii* (r = 0.13, FDR P <0.05) (**Figure 4B**). Indolepropionate was also correlated with several modules associated with cognition, such as eg. Pheynylalanine biosynthesis chorismate phenylalanine (r = 0.17, p<0.05), (**Figure 4C**).

## DISCUSSION

To our knowledge, this is the first, largest, and most comprehensive multi-omic study of the gut microbiome in a Hispanic/Latino cohort to date. Overall, our findings are consistent with prior reports of a pro-inflammatory, pathogen-enriched microbiome in cognitive decline and AD/ADRD[32, 33, 41, 45, 86]. In our MaAsLin2 regression models, abundance of anti-inflammatory, strictly anaerobic taxa, such as *Eubacterium*, e.g.: *E.eligens*, *E. siraeum*, and *E.ventriosum* was associated with better cognition. We noted an inverse association between the abundance of the sulfate-producing pathobiont *Desulfovibrio* and cognitive function, this association has been seen in prior animal and human studies[51]. Notable also, is the relationship between the identified taxa, and KEGG modules to serum metabolites, particularly BCAA and tryptophan.

In this study, higher abundance of *Alistripes obesi* and *Alistripes shahii* were associated with better cognitive function. Several *Alistipes* spp., including *Alistipes shahii* can hydrolyze tryptophan to indole[87]. Tryptophan is an essential amino acid, metabolized via host/kynurenine pathway (kynurenate, xanthurenate and quinolinate), and by gut bacteria into indole derivatives (indoleacetate, indolelactate, and indolepropionate). In prior work within HCHS/SOL-INCA, metabolites in the tryptophan pathway, notably quinolinate and kynurenine were associated with better cognitive function[88]. *Clostridium* species, including *C. phoceensis*, *C. bacterium UC511D1* and Clostridium sp KLE 1755 were also associated with better cognitive function in our study. Clostridia are known to possess genetic capacity to metabolize tryptophan[89]. Indolepropionate, a beneficial microbial tryptophan metabolite, was among the strongest correlated with several cognition-associated taxa and KEGG modules identified in this study, potentially implicating microbial tryptophan metabolism and indoles in cognition and AD/ADRD. Indolepropionate has previously been proposed to have neuroprotective properties[90] and to be of benefit as a potential therapeutic for neurodegenerative disease.[91]

Abundance of *Bacteroides cellulosilyticus*, *Bacteroides plebeius* and *Bacteroides barnesiae* were associated with better cognitive function in this study. Several Bacteroides species are involved in microbiota-derived γ-aminobutyric acid GABA[92], due to carriage a key enzyme a pyridoxal-5’-phosphate-dependent GAD (*gadB*) for converting glutamate to GABA in Bacteroides genomes. Prior work reported mixed associations between Bacteroides abundance and AD, with some studies reporting increased abundance in AD[45] and MCI[93], and others decreased abundance[94]. *Bacteroides* are gram-negative bacteria, that produce Lipopolysaccharide (LPS) as part of their cell wall[42]. LPS has been shown to lead to microglial priming, increased production of proinflammatory cytokines, neuroinflammation and neurodegeneration[43]. Bacteroides have been reported as elevated in persons with diabetes[95]. Thus, this study adds to the mixed evidence regarding Bacteroides and cognition that should be examined in future work. Two *Prevotella* species, *Prevotella sp P476* and *Prevotella bryantii* were associated with worse cognitive function in our study. Prior work on Prevotella in AD have been mixed. Lower *Prevotella* abundance has been reported in patients with MCI[96] in one study, while another study reported enrichment of *Prevotella* in AD[97]. *Prevotella* abundance has been associated with risk of Parkinson’s disease, another neurodegenerative disease[98]. Interestingly, in HCHS/SOL, a higher *Prevotella* to *Bacteroides* ratio was associated with obesity [28], while the opposite association had been reported in other populations[79].

*Bifidobacterium longum* was associated with worse cognitive function in our study. This result is somewhat surprising, given that *Bifidobacteria* are generally considered to be anti-inflammatory and beneficial to health, promoting gut epithelium integrity[99] and that *B. longum* wsa negatively correlated to most metabolites in the BCAA pathway, and strongly positively correlated with indolepropionate, a beneficial microbial metabolite, that has also been associated with reduced diabetes risk[85]. Other taxa associated with worse cognition in SOL, exhibited the opposite pattern vis. a vis these metabolites. Most, but not all prior studies have noted reduced *Bifidobacterium* abundance in AD.[45] However, increased abundance of *Bifidobacterium*[100–108] has been observed in Parkinson’s disease, including in recent work by out group[109]. Furthermore, *Bifidobacteria* abundance has been associated with aging and have been shown to be elevated in centenarians[110]. Interestingly, and more consistent with a protective role for *Bifidobacterium*, another *Bifidobacterium* species, *B. dentium* was the top species contributing to the discrimination between participants with better vs. worse cognition, with a positive (beneficial) variable importance. *Bifidobacteria* are in involved in GABA, and acetate production, and *B. dentium* is thought to be most prolific GABA producer among *Bifidobacteria*[111]. These mixed results highlight the need to further examine the role of *Bifidobacteriua* in AD/ADRD. Similarly, abundance of *R. faecalis* was associated with worse cognitive function in our study. *Ruminococcus* are generally beneficial bacteria[112], so the inverse association with cognition is surprising.

We noted dysregulation in KEGG pathways involving metabolism of neuroactive molecules (eg. GABA, ornithine), sugars (eg. ribose and glucose) and BCAA with global cognition. The GMB has a known capacity to produce as well as metabolize neurotransmitters, influencing neurotransmitter levels in the brain.[113] Several bacterial species, including members of *Bifidobacterium Bacteroides and Lactobacillus* are capable of GABA production.[114] The GABAergic system has been proposed as a potential therapeutic target for AD.[115, 116] Ornithine, derived from L-arginine and glutamate, has been implicated in the pathogenesis of AD.[117, 118] Shikimate, a tryptophan metabolism pathway, was identified as one of the pathways associated with cognition in our study, and has been linked with neurodegeneration and Alzheimer’s disease.[119–121]. This KEGG module correlated positively with tryptophan metabolite concentration in our sample. Serum tryptophan was correlated with many of the KEGG modules identified in this study, positively with modules associated with better cognition and negatively with KEGG modules associated with worse cognition. We observed correlations between other metabolites involved in tryptophan metabolism, such as the *B. longum* and indolepropionate as well as *B. cellulosityticus* and kyturenate and 3-indoxyl sulfate, suggests a potential role for microbial tryptophan metabolism in AD/ADRD.

Our study has several notable strengths. To our knowledge this is the largest and the first multi-omic study of the microbiome in Hispanics/Latinos to date. HCHS/SOL contained a culturally appropriate, comprehensive assessment of cognitive function, incorporating multiple cognitive domains. Simultaneously collected data on GMB composition, function (KEGG modules) and serum metabolites in HCHS/SOL, allowed us to conduct our multi-omic analyses, examining the inter-relationships between these features. The study also has several limitations. Because the microbiome and cognition were, at the time of writing, were only concurrently available at visit 2 in HCHS/SOL, our study was limited to a cross sectional design. Future work will consider the longitudinal relationship between GMB and cognitive decline and AD/ADRD risk. While the stool samples in this study were self-collected by the participants and based on a single collection, extensive work has shown that that self-collection is effective the microbiome is very stable with between-person variability greatly exceeding within-person differences over time.[122–124]. HCHS/SOL-INCA participants are relatively young (mean age = 60.83),. however they also have a high burden of AD-related comorbidities, such as a prevalence of diabetes or prediabetes over 80%, increasing their AD/ADRD risk beyond that appropriate for their calendar age.

In summary, our work in this large, well characterized Hispanic/Latino cohort points to a subtle, but noticeable dysregulation of the gut microbiome associated with cognitive function. We confirmed prior reports of a positive association between anti-inflammatory taxa, and negative association with pathogenic taxa and cognitive function. We also identified several KEGG pathways, with global cognitive function, which correlated strongly with serum tryptophan levels.

## SUPPLEMENTAL FIGURE LEGENDS

**Supplemental Figure 1.** Correlations within a. Taxa and b. KEGG modules associated with cognition in HCHC/SOL-INCA. Spearman (unadjusted correlations) within the top **Supplemental Figure 1A:** Taxa and **Supplemental Figure 1B:** KEGG modules associated with cognition are. Numbers in cells denote correlation coefficients, ‘*’ represents correlation p<0.05.

**Supplemental Figure 2.** Volcano Plot for MaAsLin2 model for taxa (**Supplemental Figure 2A**) and KEGG modules (**Supplemental Figure 2B**) and with additional adjustment for pack years smoking, chronic stress, depression (CESD10), hypertension, waist-hip ratio, alcohol intake, whether participant was US born, and the Multi-Ethnic Study of Atherosclerosis MESA acculturation score[83]

## ACKNOLEDGEMENTS

The authors thank the Hispanic Community Health Study/Study of Latinos- Participants for their participation in the study.

## CONFLICTS

Curtis Huttenhower serves on the Scientific Advisory Board for Seres Therapeutics and Empress Therapeutics. Tomasz Kosciolek serves as a scientific adviser to Human Biome Institute.

Other co-authors have nothing to disclose.

## FUNDING

The content is solely the responsibility of the authors and does not necessarily represent the official views of the National Institutes of Health. The Hispanic Community Health Study/Study of Latinos is a collaborative study supported by contracts from the National Heart, Lung, and Blood Institute (NHLBI) to the University of North Carolina (HHSN268201300001I / N01-HC-65233), University of Miami (HHSN268201300004I / N01-HC-65234), Albert Einstein College of Medicine (HHSN268201300002I / N01-HC-65235), University of Illinois at Chicago (HHSN268201300003I / N01-HC-65236 Northwestern Univ), and San Diego State University (HHSN268201300005I / N01-HC-65237). The following Institutes/Centers/Offices have contributed to the HCHS/SOL through a transfer of funds to the NHLBI: National Institute on Minority Health and Health Disparities,

National Institute on Deafness and Other Communication Disorders, National Institute of Dental and Craniofacial Research, National Institute of Diabetes and Digestive and Kidney Diseases, National Institute of Neurological Disorders and Stroke, NIH Institution-Office of Dietary Supplements.

This work was additionally supported by RF1AG075922, R01NS097723, 1R01MD011389, RF1AG054548, 1R01DK119268, 1R01AG048642, 1R01DK134672, R01 AG075758.

## DATA AVAILABILITY

HCHS/SOL data are archived in the dbGap and BIOLINCC managed by the National Institutes of Health. Members of the scientific community can apply to access participant data and materials at HCHS/SOL via an established process, such requests are reviewed by the project’s Steering Committee, described at https://sites.cscc.unc.edu/hchs/ (accessioned February 20, 2024). Reasonable requests for data and specimen access can be sent to the corresponding author, which will be referred to the Steering Committee of HCHS/SOL.

## CONCENT STATEMENT

all human subjects provided informed consent.

## Highlights

- Largest metagenomic study in a Hispanic/Latino cohort to date.
- *Eubacterium* species (*E. siraeum* and *E. eligens*), were associated with better cognition.
- Several KEGG modules, most strongly Ornithine, Serine biosynthesis and Urea Cycle, were associated with worse cognition.

## Research in Context

1. <u>Systematic review:</u> The authors reviewed the literature using PubMed sources. According to the literature, US Hispanics/Latinos are at an increased risk of AD/ADRD and accelerated cognitive decline compared to non-Hispanic/Latino whites and have a distinct microbial profile. While the role of the microbiome in cognitive decline and AD/ADRD has not been fully studied, and no studies have been conducted in Hispanics/Latinos to date. The relevant citations are appropriately cited in the manuscript.
2. <u>Interpretation:</u> Our study identified several species and KEGG pathways associated with cognitive function, in the largest study on Hispanics/Latinos to date. We confirmed prior reports of a positive association between anti-inflammatory taxa, and negative association with pathogenic taxa and cognitive function. We identified several KEGG modules associated with global cognitive function. Many of the cognition-associated species and functions correlated with serum tryptophan levels.
3. <u>Future directions:</u> Future, prospective work is needed to identify microbiome alterations associated with cognitive decline and dementia. More work is needed in underserve populations, such as Hispanics/Latinos.

## REFERENCES

1. Round, J.L. and S.K. Mazmanian, The gut microbiota shapes intestinal immune responses during health and disease. Nat Rev Immunol, 2009. 9(5): p. 313–23.

2. Hooper, L.V., T. Midtvedt, and J.I. Gordon, How host-microbial interactions shape the nutrient environment of the mammalian intestine. Annu Rev Nutr, 2002. 22: p. 283–307.

3. Sansonetti, P.J. and R. Medzhitov, Learning tolerance while fighting ignorance. Cell, 2009. 138(3): p. 416–20.

4. Backhed, F., et al., The gut microbiota as an environmental factor that regulates fat storage. Proc Natl Acad Sci U S A, 2004. 101(44): p. 15718–23.

5. Bercik, P., S.M. Collins, and E.F. Verdu, Microbes and the gut-brain axis. Neurogastroenterol Motil, 2012. 24(5): p. 405–13.

6. Wiertsema, S.P., et al., The Interplay between the Gut Microbiome and the Immune System in the Context of Infectious Diseases throughout Life and the Role of Nutrition in Optimizing Treatment Strategies. Nutrients, 2021. 13(3).

7. Vighi, G., et al., Allergy and the gastrointestinal system. Clin Exp Immunol, 2008. 153 Suppl 1(Suppl 1): p. 3–6.

8. Ghaisas, S., J. Maher, and A. Kanthasamy, Gut microbiome in health and disease: Linking the microbiome-gut-brain axis and environmental factors in the pathogenesis of systemic and neurodegenerative diseases. Pharmacol Ther, 2016. 158: p. 52–62.

9. Tremlett, H., et al., The gut microbiome in human neurological disease: A review. Ann Neurol, 2017. 81(3): p. 369–382.

10. Macpherson, A.J. and N.L. Harris, Interactions between commensal intestinal bacteria and the immune system. Nat Rev Immunol, 2004. 4(6): p. 478–85.

11. Gonzalez, H.M., et al., Neurocognitive function among middle-aged and older Hispanic/Latinos: results from the Hispanic Community Health Study/Study of Latinos. Arch Clin Neuropsychol, 2015. 30(1): p. 68–77.

12. Tang, M.X., et al., The APOE-epsilon4 allele and the risk of Alzheimer disease among African Americans, whites, and Hispanics. Jama, 1998. 279(10): p. 751–5.

13. Mehta, K.M. and G.W. Yeo, Systematic review of dementia prevalence and incidence in United States race/ethnic populations. Alzheimers Dement, 2017. 13(1): p. 72–83.

14. Vega, I.E., et al., Alzheimer’s Disease in the Latino Community: Intersection of Genetics and Social Determinants of Health. J Alzheimers Dis, 2017. 58(4): p. 979–992.

15. Dominguez, K., et al., Vital signs: leading causes of death, prevalence of diseases and risk factors, and use of health services among Hispanics in the United States - 2009-2013. MMWR Morb Mortal Wkly Rep, 2015. 64(17): p. 469–78.

16. Colby SL, O.J., Population Estimates and Projections. Current Population Reports. 2015, US Census Bureau. p. 25– 1143.

17. Chin, A.L., S. Negash, and R. Hamilton, Diversity and disparity in dementia: the impact of ethnoracial differences in Alzheimer disease. Alzheimer Dis Assoc Disord, 2011. 25(3): p. 187–95.

18. Luo, H., G. Yu, and B. Wu, Self-Reported Cognitive Impairment Across Racial/Ethnic Groups in the United States, National Health Interview Survey, 1997-2015. Prev Chronic Dis, 2018. 15: p. E06.

19. Fitzpatrick, A.L., et al., Sociodemographic Correlates of Cognition in the Multi-Ethnic Study of Atherosclerosis (MESA). Am J Geriatr Psychiatry, 2015. 23(7): p. 684–97.

20. Gonzalez, H.M., et al., Life’s Simple 7’s Cardiovascular Health Metrics are Associated with Hispanic/Latino Neurocognitive Function: HCHS/SOL Results. J Alzheimers Dis, 2016. 53(3): p. 955–65.

21. Almahmoud, M.F., et al., Association of Cardiac Structure and Function With Neurocognition in Hispanics/Latinos: The Echocardiographic Study of Latinos. Mayo Clin Proc Innov Qual Outcomes, 2018. 2(2): p. 165–175.

22. Tarraf, W., et al., Blood Pressure and Hispanic/Latino Cognitive Function: Hispanic Community Health Study/Study of Latinos Results. J Alzheimers Dis, 2017. 59(1): p. 31–42.

23. Lamar, M., et al., Cognitive Associates of Current and More Intensive Control of Hypertension: Findings From the Hispanic Community Health Study/Study of Latinos. Am J Hypertens, 2017. 30(6): p. 624–631.

24. Lamar, M., et al., Cardiovascular disease risk factor burden and cognition: Implications of ethnic diversity within the Hispanic Community Health Study/Study of Latinos. PLoS One, 2019. 14(4): p. e0215378.

25. Muñoz, E., et al., Stress Is Associated With Neurocognitive Function in Hispanic/Latino Adults: Results From HCHS/SOL Socio-Cultural Ancillary Study. J Gerontol B Psychol Sci Soc Sci, 2021. 76(4): p. e122–e128.

26. González, H.M., et al., *Diabetes, Cognitive Decline,* and Mild Cognitive Impairment Among Diverse Hispanics/Latinos: Study of Latinos-Investigation of Neurocognitive Aging Results (HCHS/SOL). Diabetes Care, 2020. 43(5): p. 1111–1117.

27. González, H.M., et al., Metabolic Syndrome and Neurocognition Among Diverse Middle-Aged and Older Hispanics/Latinos: HCHS/SOL Results. Diabetes Care, 2018. 41(7): p. 1501–1509.

28. Kaplan, R.C., et al., Gut microbiome composition in the Hispanic Community Health Study/Study of Latinos is shaped by geographic relocation, environmental factors, and obesity. Genome Biol, 2019. 20(1): p. 219.

29. Heston, M.B., et al., Gut inflammation associated with age and Alzheimer’s disease pathology: a human cohort study. Sci Rep, 2023. 13(1): p. 18924.

30. Verdi, S., et al., An Investigation Into Physical Frailty as a Link Between the Gut Microbiome and Cognitive Health. Front Aging Neurosci, 2018. 10: p. 398.

31. Cattaneo, A., et al., Association of brain amyloidosis with pro-inflammatory gut bacterial taxa and peripheral inflammation markers in cognitively impaired elderly. Neurobiol Aging, 2017. 49: p. 60–68.

32. Sheng, C., et al., Combination of gut microbiota and plasma amyloid-β as a potential index for identifying preclinical Alzheimer’s disease: a cross-sectional analysis from the SILCODE study. Alzheimers Res Ther, 2022. 14(1): p. 35.

33. Sheng, C., et al., Altered Gut Microbiota in Adults with Subjective Cognitive Decline: The SILCODE Study. J Alzheimers Dis, 2021. 82(2): p. 513–526.

34. Miri, S., et al., Neuromicrobiology, an emerging neurometabolic facet of the gut microbiome? Front Microbiol, 2023. 14: p. 1098412.

35. Cox, L.M. and H.L. Weiner, Microbiota Signaling Pathways that Influence Neurologic Disease. Neurotherapeutics, 2018. 15(1): p. 135–145.

36. Wiley, N.C., et al., Production of Psychoactive Metabolites by Gut Bacteria. Mod Trends Psychiatry, 2021. 32: p. 74–99.

37. Friedland, R.P., Mechanisms of molecular mimicry involving the microbiota in neurodegeneration. J Alzheimers Dis, 2015. 45(2): p. 349–62.

38. Friedland, R.P. and M.R. Chapman, The role of microbial amyloid in neurodegeneration. PLoS Pathog, 2017. 13(12): p. e1006654.

39. Zhao, Y., et al., Lipopolysaccharides (LPSs) as Potent Neurotoxic Glycolipids in Alzheimer’s Disease (AD). Int J Mol Sci, 2022. 23(20).

40. Zhao, Y. and W.J. Lukiw, Bacteroidetes Neurotoxins and Inflammatory Neurodegeneration. Mol Neurobiol, 2018. 55(12): p. 9100–9107.

41. Cattaneo, A., et al., Association of brain amyloidosis with pro-inflammatory gut bacterial taxa and peripheral inflammation markers in cognitively impaired elderly. Neurobiol Aging, 2017. 49: p. 60–68.

42. Bateman, R.J., et al., Clinical and biomarker changes in dominantly inherited Alzheimer’s disease. N Engl J Med, 2012. 367(9): p. 795–804.

43. Perry, V.H. and C. Holmes, Microglial priming in neurodegenerative disease. Nat Rev Neurol, 2014. 10(4): p. 217–24.

44. Manderino, L., et al., Preliminary Evidence for an Association Between the Composition of the Gut Microbiome and Cognitive Function in Neurologically Healthy Older Adults. J Int Neuropsychol Soc, 2017: p. 1–6.

45. Vogt, N.M., et al., Gut microbiome alterations in Alzheimer’s disease. Sci Rep, 2017. 7(1): p. 13537.

46. Haran, J.P., et al., Alzheimer’s Disease Microbiome Is Associated with Dysregulation of the Anti-Inflammatory P-Glycoprotein Pathway. mBio, 2019. 10(3).

47. Liang, X., et al., Gut microbiome, cognitive function and brain structure: a multi-omics integration analysis. Transl Neurodegener, 2022. 11(1): p. 49.

48. Zhang, L., et al., Altered Gut Microbiota in a Mouse Model of Alzheimer’s Disease. J Alzheimers Dis, 2017. 60(4): p. 1241–1257.

49. Harach, T., et al., Reduction of Abeta amyloid pathology in APPPS1 transgenic mice in the absence of gut microbiota. Sci Rep, 2017. 7: p. 41802.

50. Minter, M.R., et al., Antibiotic-induced perturbations in microbial diversity during post-natal development alters amyloid pathology in an aged APP(SWE)/PS1(ΔE9) murine model of Alzheimer’s disease. Sci Rep, 2017. 7(1): p. 10411.

51. Grabrucker, S., et al., Microbiota from Alzheimer’s patients induce deficits in cognition and hippocampal neurogenesis. Brain, 2023.

52. Ling, Z., et al., Structural and Functional Dysbiosis of Fecal Microbiota in Chinese Patients With Alzheimer’s Disease. Front Cell Dev Biol, 2020. 8: p. 634069.

53. Sawin, E.A., et al., Glycomacropeptide is a prebiotic that reduces Desulfovibrio bacteria, increases cecal short-chain fatty acids, and is anti-inflammatory in mice. Am J Physiol Gastrointest Liver Physiol, 2015. 309(7): p. G590–601.

54. Lavange, L.M., et al., Sample design and cohort selection in the Hispanic Community Health Study/Study of Latinos. Ann Epidemiol, 2010. 20(8): p. 642–9.

55. Sorlie, P.D., et al., Design and implementation of the Hispanic Community Health Study/Study of Latinos. Ann Epidemiol, 2010. 20(8): p. 629–41.

56. Callahan, C.M., et al., Six-item screener to identify cognitive impairment among potential subjects for clinical research. Med Care, 2002. 40(9): p. 771–81.

57. González, H.M., et al., A new verbal learning and memory test for English- and Spanish-speaking older people. J Int Neuropsychol Soc, 2001. 7(5): p. 544–55.

58. Lezak, M.D., et al., *Neuropsychological assessment*. 2004: Oxford University Press, USA.

59. Wechsler, D., *WAIS-R manual: Wechsler adult intelligence scale-revised*. 1981: Psychological Corporation.

60. Horn, J.L. and R.B. Cattell, Refinement and test of the theory of fluid and crystallized general intelligences. J Educ Psychol, 1966. 57(5): p. 253–70.

61. Farias, S.T., et al., The measurement of everyday cognition (ECog): scale development and psychometric properties. Neuropsychology, 2008. 22(4): p. 531–44.

62. González, H.M., et al., A research framework for cognitive aging and Alzheimer’s disease among diverse US Latinos: Design and implementation of the Hispanic Community Health Study/Study of Latinos-Investigation of Neurocognitive Aging (SOL-INCA). Alzheimers Dement, 2019. 15(12): p. 1624–1632.

63. Hillmann, B., et al., Evaluating the Information Content of Shallow Shotgun Metagenomics. mSystems, 2018. 3(6).

64. Thompson, L.R., et al., A communal catalogue reveals Earth’s multiscale microbial diversity. Nature, 2017. 551(7681): p. 457–463.

65. Marotz, C., et al., DNA extraction for streamlined metagenomics of diverse environmental samples. Biotechniques, 2017. 62(6): p. 290–293.

66. Glenn, T.C., et al., Adapterama I: universal stubs and primers for 384 unique dual-indexed or 147,456 combinatorially-indexed Illumina libraries (iTru & iNext). PeerJ, 2019. 7: p. e7755.

67. Costello, M., et al., Characterization and remediation of sample index swaps by non-redundant dual indexing on massively parallel sequencing platforms. BMC Genomics, 2018. 19(1): p. 332.

68. O’Leary, N.A., et al., Reference sequence (RefSeq) database at NCBI: current status, taxonomic expansion, and functional annotation. Nucleic Acids Res, 2016. 44(D1): p. D733–45.

69. Langmead, B. and S.L. Salzberg, Fast gapped-read alignment with Bowtie 2. Nat Methods, 2012. 9(4): p. 357–9.

70. McMurdie, P.J. and S. Holmes, *phyloseq: an R package for reproducible interactive analysis and graphics of microbiome census data*. PLoS One, 2013. 8(4): p. e61217.

71. Oksanen J, B.F., Kindt R, Legendre P, O’Hara R, Simpson GL, Solymos P, Stevens H, Wagner HH., Multivariate analysis of ecological communities in R: vegan tutorial. R package version 1.*7*.. 2013.

72. Kanehisa, M., et al., KEGG as a reference resource for gene and protein annotation. Nucleic Acids Res, 2016. 44(D1): p. D457–62.

73. Hillmann, B., et al., SHOGUN: a modular, accurate and scalable framework for microbiome quantification. Bioinformatics, 2020. 36(13): p. 4088–4090.

74. Evans, A., et al., High Resolution Mass Spectrometry Improves Data Quantity and Quality as Compared to Unit Mass Resolution Mass Spectrometry in High-Throughput Profiling Metabolomics. Metabolomics, 2004. 4(132).

75. Pernicova, I. and M. Korbonits, Metformin--mode of action and clinical implications for diabetes and cancer. Nat Rev Endocrinol, 2014. 10(3): p. 143–56.

76. Association, A.D., Diagnosis and classification of diabetes mellitus. Diabetes Care, 2013. 36 Suppl 1(Suppl 1): p. S67–74.

77. Zhernakova, A., et al., Population-based metagenomics analysis reveals markers for gut microbiome composition and diversity. Science, 2016. 352(6285): p. 565–9.

78. Peters, B.A., et al., US nativity and dietary acculturation impact the gut microbiome in a diverse US population. Isme j, 2020. 14(7): p. 1639–1650.

79. Vangay, P., et al., US Immigration Westernizes the Human Gut Microbiome. Cell, 2018. 175(4): p. 962–972.e10.

80. Siega-Riz, A.M., et al., Food-group and nutrient-density intakes by Hispanic and Latino backgrounds in the Hispanic Community Health Study/Study of Latinos. Am J Clin Nutr, 2014. 99(6): p. 1487–98.

81. Forslund, K., et al., Disentangling type 2 diabetes and metformin treatment signatures in the human gut microbiota. Nature, 2015. 528(7581): p. 262–266.

82. Mallick H., R.A., McIver L., Ma S., Zhang Y., Nguyen LH., Tickle TL., Ren B., Schawager E.H., Chatterjee S., Thompson KN., Wilkinson J.E., Subramanian Y., Lu Y., Waldron L., Paulson J.N., Franzosa EA., Bravo HC., Huttenhower C, Miltivariable Association Discovery in Population-scale Meta-omics Studies. PLos Comp Bio, 2021. In Press.

83. Kandula, N.R., et al., Association of acculturation levels and prevalence of diabetes in the multi-ethnic study of atherosclerosis (MESA). Diabetes Care, 2008. 31(8): p. 1621–8.

84. Mallick H, M.L., Rahnavard A, MA S, Zhang Y, Nguyen LH, et al., Multivariable association discovery in population-sclae meta-omics studies. 2020.

85. Qi, Q., et al., Host and gut microbial tryptophan metabolism and type 2 diabetes: an integrative analysis of host genetics, diet, gut microbiome and circulating metabolites in cohort studies. Gut, 2022. 71(6): p. 1095–1105.

86. Manderino, L., et al., Preliminary Evidence for an Association Between the Composition of the Gut Microbiome and Cognitive Function in Neurologically Healthy Older Adults. J Int Neuropsychol Soc, 2017. 23(8): p. 700–705.

87. Parker, B.J., et al., The Genus Alistipes: Gut Bacteria With Emerging Implications to Inflammation, Cancer, and Mental Health. Front Immunol, 2020. 11: p. 906.

88. He, S., et al., Blood metabolites predicting mild cognitive impairment in the study of Latinos-investigation of neurocognitive aging (HCHS/SOL). Alzheimers Dement (Amst), 2022. 14(1): p. e12259.

89. Merino, E., R.A. Jensen, and C. Yanofsky, Evolution of bacterial trp operons and their regulation. Curr Opin Microbiol, 2008. 11(2): p. 78–86.

90. Serger, E., et al., The gut metabolite indole-3 propionate promotes nerve regeneration and repair. Nature, 2022. 607(7919): p. 585–592.

91. Zhou, Y., et al., The role of the indoles in microbiota-gut-brain axis and potential therapeutic targets: A focus on human neurological and neuropsychiatric diseases. Neuropharmacology, 2023. 239: p. 109690.

92. Otaru, N., et al., GABA Production by Human Intestinal Bacteroides spp.: Prevalence, Regulation, and Role in Acid Stress Tolerance. Front Microbiol, 2021. 12: p. 656895.

93. Saji, N., et al., The relationship between the gut microbiome and mild cognitive impairment in patients without dementia: a cross-sectional study conducted in Japan. Sci Rep, 2019. 9(1): p. 19227.

94. Saji, N., et al., Analysis of the relationship between the gut microbiome and dementia: a cross-sectional study conducted in Japan. Sci Rep, 2019. 9(1): p. 1008.

95. Larsen, N., et al., Gut microbiota in human adults with type 2 diabetes differs from non-diabetic adults. PLoS One, 2010. 5(2): p. e9085.

96. Kim, E.J., et al., Association between Mild Cognitive Impairment and Gut Microbiota in Elderly Korean Patients. J Microbiol Biotechnol, 2023. 33(10): p. 1376–1383.

97. Kaiyrlykyzy, A., et al., Study of gut microbiota alterations in Alzheimer’s dementia patients from Kazakhstan. Sci Rep, 2022. 12(1): p. 15115.

98. Li, Z., et al., Gut bacterial profiles in Parkinson’s disease: A systematic review. CNS Neurosci Ther, 2023. 29(1): p. 140–157.

99. Wang, Z., et al., The role of bifidobacteria in gut barrier function after thermal injury in rats. J Trauma, 2006. 61(3): p. 650–7.

100. Aho, V.T.E., et al., Gut microbiota in Parkinson’s disease: Temporal stability and relations to disease progression. EBioMedicine, 2019. 44: p. 691–707.

101. Bolliri, C., et al., Gut Microbiota in Monozygotic Twins Discordant for Parkinson’s Disease. Ann Neurol, 2022. 92(4): p. 631–636.

102. Wallen, Z.D., et al., Metagenomics of Parkinson’s disease implicates the gut microbiome in multiple disease mechanisms. Nat Commun, 2022. 13(1): p. 6958.

103. Lin, A., et al., Gut microbiota in patients with Parkinson’s disease in southern China. Parkinsonism Relat Disord, 2018. 53: p. 82–88.

104. Li, Y., et al., [Features of gut microbiota in patients with idiopathic Parkinson’s disease]. Zhonghua Yi Xue Za Zhi, 2020. 100(13): p. 1017–1022.

105. Unger, M.M., et al., Short chain fatty acids and gut microbiota differ between patients with Parkinson’s disease and age-matched controls. Parkinsonism Relat Disord, 2016. 32: p. 66–72.

106. Barichella, M., et al., Unraveling gut microbiota in Parkinson’s disease and atypical parkinsonism. Mov Disord, 2019. 34(3): p. 396–405.

107. Hill-Burns, E.M., et al., Parkinson’s disease and Parkinson’s disease medications have distinct signatures of the gut microbiome. Mov Disord, 2017. 32(5): p. 739–749.

108. Petrov, V.A., et al., Analysis of Gut Microbiota in Patients with Parkinson’s Disease. Bull Exp Biol Med, 2017. 162(6): p. 734–737.

109. Palacios, N., et al., Metagenomics of the Gut Microbiome in Parkinson’s Disease: Prodromal Changes. Ann Neurol, 2023. 94(3): p. 486–501.

110. Palmas, V., et al., Gut Microbiota Markers and Dietary Habits Associated with Extreme Longevity in Healthy Sardinian Centenarians. Nutrients, 2022. 14(12).

111. Barrett, E., et al., γ-Aminobutyric acid production by culturable bacteria from the human intestine. J Appl Microbiol, 2012. 113(2): p. 411–7.

112. La Reau, A.J. and G. Suen, The Ruminococci: key symbionts of the gut ecosystem. J Microbiol, 2018. 56(3): p. 199–208.

113. Olson, C.A., et al., The Gut Microbiota Mediates the Anti-Seizure Effects of the Ketogenic Diet. Cell, 2018. 173(7): p. 1728–1741.e13.

114. Bravo, J.A., et al., Ingestion of Lactobacillus strain regulates emotional behavior and central GABA receptor expression in a mouse via the vagus nerve. Proc Natl Acad Sci U S A, 2011. 108(38): p. 16050–5.

115. Calvo-Flores Guzmán, B., et al., The GABAergic system as a therapeutic target for Alzheimer’s disease. J Neurochem, 2018. 146(6): p. 649–669.

116. Govindpani, K., et al., Towards a Better Understanding of GABAergic Remodeling in Alzheimer’s Disease. Int J Mol Sci, 2017. 18(8).

117. Liu, P., et al., Altered arginine metabolism in Alzheimer’s disease brains. Neurobiol Aging, 2014. 35(9): p. 1992–2003.

118. Liu, P., Y. Jing, and H. Zhang, Age-related changes in arginine and its metabolites in memory-associated brain structures. Neuroscience, 2009. 164(2): p. 611–28.

119. Paley, E.L., Diet-Related Metabolic Perturbations of Gut Microbial Shikimate Pathway-Tryptamine-tRNA Aminoacylation-Protein Synthesis in Human Health and Disease. Int J Tryptophan Res, 2019. 12: p. 1178646919834550.

120. Paley, E.L., et al., Geographical Distribution and Diversity of Gut Microbial NADH:Ubiquinone Oxidoreductase Sequence Associated with Alzheimer’s Disease. J Alzheimers Dis, 2018. 61(4): p. 1531–1540.

121. Paley, E.L. and G. Perry, Towards an Integrative Understanding of tRNA Aminoacylation-Diet-Host-Gut Microbiome Interactions in Neurodegeneration. Nutrients, 2018. 10(4).

122. Faith, J.J., et al., The long-term stability of the human gut microbiota. Science, 2013. 341(6141): p. 1237439.

123. Franzosa, E.A., et al., Identifying personal microbiomes using metagenomic codes. Proc Natl Acad Sci U S A, 2015. 112(22): p. E2930–8.

124. Mehta, R.S., et al., Stability of the human faecal microbiome in a cohort of adult men. Nat Microbiol, 2018. 3(3): p. 347–355.

125. Mirzayi, C., et al., Reporting guidelines for human microbiome research: the STORMS checklist. Nat Med, 2021. 27(11): p. 1885–1892.

